# A protocol for a systematic critical realist synthesis of school mindfulness interventions designed to promote pupils’ mental wellbeing

**DOI:** 10.1101/2023.04.06.23288255

**Authors:** Pamela Abbott, Graeme Nixon, Isabel Stanley, Lucia D’Ambruoso

**Affiliations:** Centre for Global Development, University of Aberdeen, Aberdeen, UK; School of Education, University of Aberdeen, Aberdeen, UK; Centre for Data Science, University of Aberdeen, Aberdeen, UK

**Keywords:** Mindfulness, schools, mental wellbeing, children and adolescents, critical realism, complexity theory

## Abstract

**Introduction:** The review described in this protocol will be the first critical realist review of the literature reporting on the impact of mindfulness interventions in schools on the mental wellbeing of pupils. Mindfulness interventions are increasingly being introduced into schools to promote children’s (and teachers’) wellbeing. Findings from impact evaluations, including systematic reviews and metanalysis, suggest that school-based mindfulness interventions promote pupils’ wellbeing. However, there is a need for further evidence on how they work and for whom and under what circumstances.

**Methods and analysis:** A critical realist review methodology will be used to provide a causal interdisciplinary understanding of how mindfulness interventions in schools promote the mental wellbeing of pupils. This will be done through a systematic literature review and extrapolating context, agency, intervention, mechanisms, and outcome configurations. This will enable us to understand how in certain contexts, pupils can use the resources offered by a mindfulness intervention to trigger mechanisms that promote their mental wellbeing and what mechanisms in the context support, restrict or prevent change. We will then use retrodiction and retroduction to develop the most plausible interdisciplinary middle-range theory to explain the findings.

**Discussion:** The review findings will inform a critical realist evaluation of a mindfulness intervention in schools that we will be carrying out. The findings from the review will enable us to inform policymakers and other stakeholders about what conditions need to be in place for mindfulness interventions to promote pupils’ mental wellbeing and for which pupils. We will publish the findings from the review in academic and professional publications, policy briefs, workshops, conferences, and social media.

## Introduction

This critical realist review aims to understand how and why school-based mindfulness interventions (SBMIs) work or do not work in promoting children’s mental wellbeing. The broad purpose is to move from empirical observation to develop theorised understanding. This will produce knowledge that enables us to make recommendations to improve practice, that is, the delivery of mindfulness interventions in schools (1). In this sense, we can locate the main purpose, to build a middle-range theory (2) that identifies the discursive formation in which it is most fruitful to discuss the phenomena (i.e. the ontology within which the results are to be understood), and uses this discourse to model the main underlying mechanisms. It proceeds to explain how they are triggered by the agency of pupils in response to the mindfulness interventions and how these interactions and interconnections make the interventions work (or not and to what extent) in given contexts (schools, neighbourhoods, countries). The findings will enable us to refine the programme theory for research we are carrying out examining the potential for SBMIs to promote the mental wellbeing of children and adolescents. The findings will also support policymakers and others globally to implement policies promoting the mental wellbeing of children and adolescents through SBMIs.

Poor mental wellbeing is a serious global challenge (3–6), with the lifetime risk of experiencing mental health problems being 28-41% in low-and-middle-income countries and 17-49% in high-income countries (6). Income and gender inequalities exacerbate the problem, with women being nearly twice as likely to develop depressive illnesses as men (7). Poor people are likelier to have mental health problems than those better off (8). It is also estimated that just under 50% of adults with mental health issues experienced them before they were 18 and a third before 14 years (9). A life-course approach is, therefore, essential to break the cycle of intergenerational transfer of mental disorders (6,10). Promoting CA mental wellbeing is even more urgent owing to the negative impacts COVID-19 has had on their mental wellbeing, including girls having been more negatively affected than boys (3,11).

All children have a right to enjoy their childhood, attain their full potential, and lead productive adult lives (12). The absence of mental wellbeing means we can envisage a ’real’ utopia (13) where the mental wellbeing of all children is promoted. However, the low prioritisation of mental wellbeing means that children are vulnerable to abuse of their human rights as set out in the United Nations Convention *on the Rights of the Child* (14). The benefits of promoting mental wellbeing to children, their families and societies are high, meaning that there is a strong economic as well as a moral case for promoting their mental wellbeing (15,16). Improved mental wellbeing improves individuals’ quality of life and productivity, and collective improvements in mental wellbeing reduce healthcare costs.

Schools are seen as an appropriate place to deliver interventions because of how much time children spend in them and the economies of scale that can be achieved (15,17,18). Such interventions benefit all children, empower them, are preventative and have some therapeutic value. The WHO recommends providing mental wellbeing interventions for all children (19,20).

Mindfulness has been identified as an effective intervention for promoting children’s mental wellbeing, as evidenced by systematic reviews and meta-analyses (21–23), especially when integrated into a whole-school approach (21,24,25). Mindfulness training is relevant for CA across the spectrum of mental wellbeing (26). SBMIs aim to support the education system by reducing stress in pupils and teachers, reducing teacher absenteeism and burnout, and improving pupil learning outcomes. Systematic reviews and meta-analyses of the impact of SBMIs have shown positive outcomes, including increased prosocial behaviour and resilience, improved attention, cognitive and socio-emotional skills, and better academic performance in general, as well as reduced stress. Mindfulness interventions also reduce teachers’ stress levels, improve their mental wellbeing and life satisfaction, and their relationships with students making classroom environments more conducive to student learning (27–31). The finding that mindfulness has a positive effect across several outcome domains has led to calls for the broad introduction of mindfulness in schools (28,31,32). The consensus is that mindfulness is unlikely to cause harm, and the potential benefits are high (33,34).

Whole-school interventions have the advantage of being relatively inexpensive, non-stigmatising and inclusive. They can be integrated into the school curriculum and taught by teachers after a short pre- or in-service training (16). Locating the intervention in schools recognises that mindfulness is fundamentally relational and only has meaning when it is a shared social practice (35,36). Whole school interventions also aim to improve the school climate, create prosocial classrooms, improve pupil-teacher and pupil-pupil relations, increase children’s engagement, and support pupil and teacher wellbeing (23,37). Such interventions aim to create schools where pupils and teachers share a disposition to mindfulness and a sense of personal control and investment and feel more in control of their lives. They enable pupils to develop a range of educational skills, including ’the flexible transfer of skills and knowledge to new contexts, the development of deep understanding, student motivation and engagement, the ability to think critically and creatively, and the development of more self-directed learners’ (38). SBMIs are then defended against the charge of commodifying children and producing neoliberal subjects because they bring about, or can bring about, educational aims and benefits (in addition to socioeconomic ones) (16,37,39,40).

Promising as these findings are, we are left with unresolved issues that limit our ability to use them. While systematic reviews have indicated promise regarding impacts, there have been varying outcomes, and not all interventions have been shown to work (41). Mindfulness-based interventions consist of several elements, vary in theoretical underpinnings, duration and mode of delivery, and have heterogeneous outcomes (16,42). Most studies are from Western, high-income countries, most notably the USA and the UK, with few in low-and-middle-income countries (LMICs). Most notably, the evidence does not indicate *what makes these programmes work, how, where, with whom and to what extent*. Researchers’ primary focus has been asking if mindfulness works rather than how and why it works (43). Most theories assume that practising mindfulness exercises bring about the observed changes (44) and that mindfulness is about changing the wiring of the brain and the psychology of individuals (39,42,44). A scoping review to elucidate the mediators, moderators and implementation factors in delivering universal mindfulness training in schools (45) identified mediators and moderators that influenced the outcome of the intervention. These included the fidelity of the intervention, the dose, the quality, the reach, and the receptivity of the pupils. However, the analysis is descriptive rather than explanatory. The authors also point out that few studies of SBMIs report findings from the process evaluation. It concluded that what is required is higher quality trials that include process evaluation.

However, constant conjunction does not explain how mindfulness works, and psychological theories of mindfulness are inadequate for explaining how an intervention designed to promote mental wellbeing, a bio-psycho-social phenomenon, works. Evidence, for example, suggests that group effects are important (46), as suggested by findings from whole-school interventions (23,37,38). Mindful schools cultivate mindfulness as a disposition that changes pupil (and teacher) behaviour, thereby improving the school ’climate’. Evidence from low, middle and high-income countries shows that school climate is important for positive student outcomes, including mental wellbeing, with feedback loops between teachers’ improved mental wellbeing and pupils’ (47–49). A realist review of MBIs at the workplace designed to improve employee welling found that the intervention triggered four mechanisms that participants were able to use: awareness/self-regulation, acceptance/compassion, feeling permitted to take care of the self, and a sense of growth and being able to attain goals (44). Critically, this research also found that a *supportive environment at work* was important, linking mindfulness practice to specific practice settings and existing goals and practices.

The fear of being seen as non-productive stimulated the adoption of mindful practices and reduced the extent of non-adherence/dropout.

Given the limitations of existing evidence on how SBMIs work, new interdisciplinary knowledge considering their net effects and investigations of how and why, for whom and under what circumstances they work (or do not) will provide the necessary evidence for policy and decision-makers. Such research can generate the knowledge to understand better what needs to be done to implement SBMIs, so they benefit pupils’ and teachers’ mental wellbeing and improve their quality of life. To do so, they must open the black box and map the complex pathways linking the mindfulness intervention to outcomes. The conclusion from recent SBMI trials is that this should be a priority for future research (50,51) rather than the usual recommendation of more trials of higher quality to demonstrate efficacy definitively. That SBMIs and improvements in child mental wellbeing are correlated in many instances has established a ’demi-regularity’. It is this demi-regularity that needs to be explained (52). We need to determine the contexts and agential responses to the mindfulness interventions that enable them to work for some children in some contexts and not others.

## Aim and Objectives

The aims of the review are to:

a. describe plausible explanations for the effectiveness of mindfulness interventions designed to promote pupils’ wellbeing, and
b. create transferable theories that can inform programme design and implementation in different settings.

To achieve the aim, the objectives are to identify:

- theories about how mindfulness interventions work in schools;
- the contexts and mechanisms that may facilitate or hinder implementation;
- how pupils and teachers respond to mindfulness interventions (agency);
- how school contexts influence the agency of pupils in responding to the mindfulness intervention and trigger mechanisms that change the context and lead to outcomes;
- how the school system changes (roles and relationships), including pupil-teacher relations and pupil-pupil-relations;
- how the school attitudes and values change (culture), and;
- the outcomes resulting from the interventions.

## Methodology

### Introduction

To achieve our aims and objectives, we will conduct a systematic critical realist review to identify how mindfulness interventions promote pupils’ mental wellbeing. A critical realist review is explanatory; it seeks to explain how interventions work and generate different outcomes in different contexts (53,54). We will explore how mindfulness interventions are supported or inhibited by contextual mechanisms in schools, how CA and teachers respond to them and the outcomes that result from the interaction between the intervention and contextual mechanisms and the response of CA and teachers. In doing so, we will identify the ’demi-regularities’, the contexts over time and space in which mindfulness interventions enable pupils’ agency to trigger mechanisms that promote their mental wellbeing (52).

Because critical realism affirms the reality of objects, agents, and mechanisms, which cannot be viewed directly but only deduced from their effects, it seeks to identify, by retroduction and retrodiction, the middle-range interdisciplinary theory that most comprehensively explains how mindfulness interventions work. Such theories are always open to refinement in the light of new evidence.

There is no agreed standard or guide for critical realist reviews, but they are underpinned by a critical realist meta-theory and draw on some elements of the realist review methodology of Roy Pawson (1). We have adapted the standard for realist reviews (55,56) and taken into account recommendations for traditional systematic reviews (57–59). PRISMA offers transparency, validity, replicability, and updateability (Additional Material 1).

### A critical realist research paradigm

Our methodology is informed by critical realist metatheory (Table 1 defines key critical realist terms used in this protocol). Critical realism in the social and health sciences is most frequently associated with the work of Roy Bhaskar (60). It has informed the development of methods for systematic reviews and impact evaluations that are designed to explain how and why interventions work in the ways they do (61). However, few systematic reviews and impact evaluations have used the approach. Understanding how meta-theory can inform carrying out and reporting on critical realist systematic reviews is still evolving.

**Table 1:** Critical Realist Terminology

|  |  |
| --- | --- |
| **Agency** | The power of individuals to engage in meaningful actions. Agents have causal powers because they respond to the intervention and, within their context, can trigger mechanisms that change the context. |
| **Abduction** | Reinterpreting events into a conceptual framework. |
| **Complexity** | Outcomes result from the complex interaction of mechanisms. Therefore, change is non-linear and involves feedback loops. |
| **Context** | The context sets limits to the efficacy of a programme intervention. Structures are material, physical, and human resources and associated practices. Culture is the realm of intersubjectivity, social values/ideas, and ideational influences. Contexts are dynamic and can change through agency triggered by the intervention. |
| **Context Mechanisms** | The resources and restrictions embedded in the social and organisational (relational) structure that can influence the outcomes and must be identified. They can be physical, social, |
|  | interpersonal, intrapersonal, or conceptual and have causal powers. While they may not be observable, they can be retrodeduced from what is observed. |
| **Culture** | Intersubjectivity, ideas and ideational influences. |
| **Demi-regularities** | Partial event regularities indicate that an intervention in some contexts triggers mechanisms that result in the same outcome. |
| **Depth Ontology** | Reality is stratified into three domains.<br>1. The Empirical: experiences of what actually happens;<br>2. The Actual: things that happen independently of whether we observe them or not;<br>3. The Real: constituted by mechanisms with generative power, events, and experiences. |
| **Emergence** | The ability of mechanisms to combine to create something new that cannot be reduced to the mechanisms from which it emerged. |
| **Intervention** | The intervention is designed to bring about change in a given context. |
| **Intervention mechanisms** | The mechanisms triggered by the experiences, interpretations, and responses to interventions by actors that can change the context mechanisms. |
| **Judgemental Rationalism** | The evaluation of competing explanations to identify the one that has the most credible explanation, the one that is most practically adequate. |
| **Laminated System** | The social world is understood as an open, complex, stratified (layered) system of objects that make things happen. There are seven layers, from the Global to the sub-individual level, each influencing the other. |
| **Middle-Range Theory** | Middle-range theories lie between untestable grand theory and concrete description. They are theories that can generate hypotheses that can be tested and refined through empirical research. |
| **Morphogenic Approach** | Based on the view that there are three primary causal powers in society, structure, culture and agency, and that structure and culture necessarily predate the actions that transform them. |
| **Open System** | A (social) system where multiple causal factors interact and influence each other, and it is impossible to isolate variables as in laboratory conditions. The multiplicity of mechanisms other than the ones triggered by the intervention means that what happens may not necessarily be what was envisaged. |
| **Outcome** | Outcomes are the changes in the context that result from how people respond to the intervention and the contextual mechanisms. |
| **Retrodiction** | Determining what mix of mechanisms interact reinforce, moderate, or counteract) to produce the outcomes. |
| **Retroduction** | Inferential thinking – what makes the observable phenomena possible, what must the real be like. |
| **Structure** | Material, physical, and human resources and their associated practices at the level of social and systems integration, that is, the relations between members of society and the relationship between institutions |
| **Transitive and Intransitive Dimensions** | The intransitive dimension is the physical and social world we inhabit; things are as they are and are not constituted by our understanding of them and are independent of how we describe them. The transitive is the theories and discourses we hold in order to understand our world. |

**Table 2:** Search Terms for IngentaConnect and Scopus

|  |  |
| --- | --- |
| **IngentaConnect:** | article for (Title, Keywords or Abstract contains 'mindfulness OR mindful OR "mindfulness-based cognitive therapy" OR MBCT OR MBSR AND school OR "whole school" OR "school-based" OR "educational context" AND children OR youth OR "young people" OR juvenile OR teen OR "young adult" OR teenager')<br>Then due to last entry not fitting, second search of:<br>article for (Title, Keywords or Abstract contains 'mindfulness OR mindful OR "mindfulness-based cognitive therapy" OR MBCT OR MBSR AND school OR "whole school" OR "school-based" OR "educational context" AND children OR youth OR "young people" OR juvenile OR teen OR "young adult" OR pupils') |
| **Scopus** | Search within Article title, Abstract, Keywords: mindfulness OR mindful OR "mindfulness-based cognitive therapy" OR MBCT OR MBSR AND school OR "whole school" OR "school-based" OR "educational context" AND children OR adolescents OR youth OR |
|  | "young people" OR juvenile OR teen OR "young adult" OR "teenager"<br>OR pupils |

Critical realism provides guidelines for inter/transdisciplinary research (13,62–64), including research on promoting wellbeing (13,65). Inter/transdisciplinary research is essential when considering mental wellbeing. It is a bio-psycho-social phenomenon and requires the identification of the biological, psychological and social generative mechanisms (65–68). The outcome of SBMIs are likely due to the complex interaction of neurobiological, cognitive, psychological, social-structural and cultural mechanisms. The interdisciplinary team must include researchers with expertise in these disciplines committed to working together to develop trans-factual middle-range theories of how the intervention works (13).

Critical realist research is evolving, and there is no single approach. Critical realism is a philosophy of science that in itself explains nothing; it is a meta-theory of how it is possible to have scientific knowledge about the social world. What is necessary is to develop an explanatory programme. Our approach draws on Margaret Archer’s practical morphogenic approach (70,71), a methodology that complements Critical Realism’s social ontology (72). We developed our approach to the literature review, drawing on Porter’s and Hind’s and Dicksons’ identifications (from the perspective of critical realism) of the weaknesses of the realist approach to systematic reviews based on the work of Ray Pawson (73–75).

Critical realism is: ontologically realist, arguing that phenomena exist in the social world relatively independent of what we know or think about them, and epistemologically interpretivist, arguing that the way we come to know about phenomena is context-dependent, fallible, prone to individual interpretation, and seen from our angle of vision (61,76). Critical realism rejects Humean’ constant conjunctions’ that B invariably follows A as an empirical basis for conceptualising social reality. They argue that correlations are descriptive and fallible and do not explain how interventions work. Instead, they argue that there are demi-regularities, and it is how they occur that needs to be explained; that is why an intervention works in some contexts and not others and for some people and not others (52). Critical realism is theory-driven and seeks to explain/theorise why things occur in the way they do, what the (generally hidden) mechanisms are that generate events, recognise that social systems are open, eliminated and layered and that events are often the outcome of the complex interaction of mechanisms (75,77–80). The effectiveness of an intervention is not based only on its inherent qualities but on the action of agents in taking up the opportunities afforded by the intervention to trigger mechanisms that result in the outcomes (60,71). Change is non-linear and often involves complex feedback loops (78,79,81,82). Critical realists are committed to social justice, moving from how things are to how they should be to improve people’s lives enabling fairness in society (83–85). Different contexts may be more or less supportive of change; therefore, outcomes are context-specific, and all theories are fallible and open to modification or refutation.

Archer’s morphogenic approach is the methodological complement to Bhaskar’s model of social transformation (71). She argues that every theory about the social involves understanding the relationship between structure, agency, and culture (SAC) (Figure 1). She argues against conflating agency with structure; the context in which we live comprises both structural and cultural mechanisms, and that these mechanisms are analytically separable from and necessarily predate the agency which reproduces or transforms them. Structural and cultural mechanisms are real; they exist independently of us and place limitations on opportunities for agency. The context in which agents live shapes their beliefs, desires, and opportunities and limits their agency – context conditioning. However, the interaction between context and agency shapes and reshapes the context; agency can change the context (morphogenesis) or reproduce it (morphostasis). There are three morphogenetic cycles, material, cultural and agency. An intervention such as introducing mindfulness into a school aims to bring about change by giving pupils and teachers the resources to trigger mechanisms that can lead to material and cultural change. Their responses to the intervention are shaped, but not determined, by the context, and individuals generate outcomes through actions and interactions. When actors trigger new mechanisms (material and/or cultural), the context changes. However, pupils and teachers can resist, redefine, repudiate, suspend, or circumvent engagement with the intervention.

**Figure 1:** Intervention Cycle Showing Context, Agency, Intervention, Mechanisms Outcome (CAIMO) Configuration

### Methods/Design

### Step 1: establishing the scope of the work

The review will focus on SBMIs designed to promote the mental wellbeing of pupils in high-, middle-and low-income countries. We will document differences between SBMIs underpinned by different theories, aims, approaches and techniques and delivered differently for different lengths of time (86,87). The review will also capture other individual differences and programme characteristics that can impact programme receptivity and impact (24). We will only include school mindfulness interventions that include mindfulness meditation as a central pedagogical component (16,88) aimed at improving pupils’ behavioural, cognitive and mental wellbeing outcomes. Nevertheless, outcomes will likely vary by the precise nature of the mindfulness intervention.

### Step 2: search for evidence

#### Search techniques

A rigorous systematic Preferred Reporting Items for Systematic Reviews and Meta-Analysis (PRISMA) approach will be used to search for literature (89). A PRISMA diagram will show the steps of the inclusion and exclusion of documents (Additional Material 2). The literature search will be in three phases: searching electronic databases, searching other sources such as relevant journals and core publishers, and citation tracking to ensure all relevant studies are included. The aim is to include as wide a range as possible of academic and grey literature without restrictions on study type or publication date. No location restrictions will be applied to gain a wide range of relevant studies internationally in high-, middle- and low-income countries. Databases that index health, psychology, sociology and/or education literature will be searched. The search will be restricted to publications in the English language. The search terms and the databases used are based on the advice of an academic librarian.

Twenty-one scholarly databases will be searched: MEDLINE, PsychoInfo, Web of Science Core Collection, PubMed Central, Google Scholar, Cochrane Library, Proquest, SciELO Citation Index, Embase, Sociological Abstracts, Scopus, Current Content Connected, Child Development and Adolescent Studies, British Education Index, Data Citation Index (Clarivate), CINAHL, ERIC, Education Abstracts, Education Research Abstracts, IngentaConnect, and JSTOR.

Seven databases will be used to search for the grey literature: the Bielefeld Academic Search Engine, OpenGrey, PsycEXTRA, ProQuest Dissertations and Theses, Grey Matters, ResearchGate, Academia.edu, and the Social Science Research Network.

For books and book chapters, the websites of eight academic publishers will be searched, Routledge (Taylor & Francis), Palgrave Macmillan, Springer, Elsevier, Wiley, SAGE, Oxford University Press and Cambridge University Press.

The Search terms will be: ’mindfulness’ or ’mindful’ or ’mindfulness-based cognitive therapy’ or MBCT or MBSR and ’school’ or ’whole school’ or ’school-based’ or ’educational context’ and ’children’ or ’adolescents’ or ’youth’ or ’young people’ or’ juvenile’ or ’teen’ or’ young adult’ or ’teenager’ or ’pupils’.

#### Inclusion/exclusion criteria Inclusion

- Study design: all published study types reporting empirical research but excluding literature reviews and meta-analysis. (The references of literature reviews and meta- analyses will be scrutinised to ensure that all studies are included in the review. We will also record their main conclusions on a separate extraction table).
- Documents: from any country or region.
- Publication date: any.
- Document type: any document type that will inform the review.
- Population: children aged 7-16 years participating in a SBMI.
- Intervention: includes mindfulness meditation as a central pedagogical component and aims to improve behavioural, cognitive, and mental wellbeing outcomes.
- Language: English.

#### Exclusion

- Only includes special needs schools.
- Therapeutic interventions targeted at pupils with mental health problems.
- Only includes meditation, with mindfulness meditation not being a central pedagogical component.
- Only includes yoga.
- Only includes pupils outside the 7-16 years age group.
- In languages other than English.

#### Article screening

*Covidence* will be used to manage article screening and data extraction.

1. Remove duplicates and citations without abstracts or summaries;
2. Two reviewers (PA and IS) will review the titles and abstracts of all retrieved documents captured by our search strategy and code them as ’potentially relevant’ and ’not relevant’. Any disagreements will be resolved by discussion or, if necessary, bringing in a third reviewer (LD);
3. Download the full text of potentially relevant documents.

### Step 3: document appraisal and data extraction

Extract information from the documents that potentially meet our inclusion criteria into an Excel spreadsheet:

1. Document details – title, authors, year of publication, location of study;
2. Country, income group (low, lower-middle, upper-middle, high) and main religion(s);
3. The mindfulness intervention description – the type of MBI, trainers, design, aim/purpose, length of training;
4. Sample characteristics- age of pupils, sex/gender, socioeconomic status, ethnicity, type of school;
5. The study design and if it is fit for purpose (quality/rigour);
6. Rational for SBMI, including any social justice framing;
7. Contextual factors (mechanisms) before the intervention was introduced;
8. Proximal outcomes measured;
9. How proximal outcomes were measured;
10. Outcomes;
11. Agency;
12. Generative mechanisms triggered by the intervention that could have supported change (positive mechanisms);
13. Generative mechanisms triggered by the intervention that could have restricted/prevented change (negative mechanisms);
14. Contextual mechanisms that could have supported change (positive mechanisms);
15. Contextual mechanisms that restricted/prevented change (negative mechanisms;
16. Any theoretical explanations identified for explaining the outcomes and the level of the explanation that is, biological, psychological, or social;
17. Changes in context following the introduction of the mindfulness intervention.

The extraction tool will be piloted. PA, LD, IS and GN will independently read two documents and complete the extraction table. They will then meet, compare their extraction tables, and agree on any necessary modifications.

PA will extract all information. PA will remove any documents containing insufficient relevant data to inform how and why the intervention worked (or did not work) and/or not using credible and trustworthy methods. Documents will be considered relevant if they can help to answer the research questions; that is, they report findings from research on SBMIs. They will be included as credible if the methods used are adequate for generating the findings; documents will be excluded if they are not based on credible research or are purely anecdotal. The reasons for the exclusion of any document will be noted. A critical realist synthesis does not require that two independent reviewers complete screening for quality or data relevance. However, LD will review any documents PA identifies as not contributing or not using credible and trustworthy methods, with differences being resolved by discussion and, if necessary, by bringing in a third reviewer, GN.

We will provide a descriptive narrative summary of the findings from this stage of the review.

### Step 5: Analysis and Reporting

The analysis will aim to identify a middle-range interdisciplinary theory that explains how mindfulness interventions work in schools to promote pupils’ mental wellbeing. It will open the black box and identify the mechanisms the mindfulness interventions triggered that explain how the intervention caused the reported outcomes (Figure 2). An interdisciplinary middle-range theory will then be developed that explains how mindfulness interventions work, recognising that outcomes will likely differ in different contexts.

**Figure 2:** Hypothetical Intervention Cycle Opening Up the Black Box

The analysis will follow the critical realist stages of analysis for applied interdisciplinary research, Resolution, Redescription, Retrodiction, Elimination, Identification, Correction (RRREIc) (Table 3) (13,90). Mental wellbeing is the outcome of a complex interaction of bio- psycho-social mechanisms. An adequate explanation of how mindfulness interventions impact children’s mental wellbeing must be interdisciplinary (65). The framework consists of six steps to build transdisciplinary accounts of phenomena. These are: (1) break down complex events into component parts and identify disciplinary explanations of impact; (2) redescribe impacts in mono-theoretically meaningful ways; (3) use retrodiction to identify trans-factual theories that explain the ways the mechanisms interact to produce outcomes; (4) eliminate alternative competing hypotheses; (5) identify the most comprehensive explanation; (6) refine scientific knowledge in light of (provisional) findings (91).

**Table 3:** RRREIc Stages for Interdisciplinary Research

| Planning Phase | Disciplinary Phase | Transdisciplinary Phase | Interdisciplinary Phase |
| --- | --- | --- | --- |

| Resolution | Redescription | Retrodiction | Elimination | Identification | Refinement |
| --- | --- | --- | --- | --- | --- |
| Identification of the levels that are included. Biological/ Individual Social- Interaction Socioeconomic | Identification of disciplines and using abduction to develop theoretical explanations for the impact of the intervention on children's mental wellbeing | Identify how different mechanisms reinforce, moderate, and condition one another to affect the outcome | Elimination of alternative theoretical explanations. | Identify the most comprehensive interdisciplinary explanation for how mindfulness promotes children's mental wellbeing |  |
| Identification of disciplines that research at different levels | Develop disciplinary theories to explain the impact. Reductionist/ atomistic explanations | Develop trans-factual theories integrating the disciplinary explanations. | Use judgemental rationality to eliminate theories that lack explanatory power | Use judgemental rationality to identify the most comprehensive explanation | Iterative refinement of theory |

To do this, we will first build CAIMOs for each mechanism identified (92). This will involve a detailed and systematic analysis based on the outcome patterns, context, agency, interventions, mechanisms triggered by agency, and outcomes: the CAIMO configuration (Figure 4). The four critical realist evaluation categories are the context before the intervention, the children’s agency, the mechanisms triggered by children’s (and teacher’s) agency and the outcomes. Agency is pupils’ (and teachers’) responses to the mindfulness interventions, mechanisms are triggered by agency and enable pupils (and teachers) to benefit from the intervention, and the pre-existing structures and culture are the contextual conditions that facilitate or inhibit pupils’ potential to benefit from the intervention (see Figure 3).

**Figure 3:** Critical Realist Evaluation Process

**Figure 4:** Simplified Hypothetical Causal Pathway of Change after an Intervention

Outcomes patterns are the mental wellbeing outcomes pupils experience from participating in SBMIs. Within each category, findings will be broken down thematically and reported narratively to distinguish between different contexts, agency responses, mechanisms triggered, and outcomes (Figure 4). The key themes that describe processes and causal mechanisms for explaining mindfulness intervention outcomes in schools will then be identified. Hypothetical links will then be made between the CAIMO themes, creating potential pathways that account for the impacts of school-based mindfulness interventions on pupils and why, for whom and under what circumstances these impacts occur.

However, to take account of complexity, it is necessary to develop a non-linear pathway of change showing how the complex interaction of mechanisms (context mechanisms that predate the intervention and those triggered by the intervention) leads to the observed outcomes (Figure 4) (75,79,80,93). To do this, we will use feedback loop diagrams to model change, showing both mechanisms triggered by the intervention that cause change, those already in the context that supported change (+ve mechanism) and those already in the context and mechanisms triggered by the intervention that restricted or prevented change (- ve mechanisms). Outcomes will likely be more complex than a dichotomy between morphogenesis (structural change) and morphostasis (structural reproduction).

### Ethics

Formal ethics approval is not required for a literature review. However, ethical approval has been obtained from the University of Aberdeen, Addis Ababa University, and the University of Rwanda for the research programme, of which the critical realist review forms an integral element.

### Dissemination

We will publish at least one article in a peer review journal reporting the findings from the literature review, conforming to RAMESES publication standards (56) and a policy brief intended for policymakers with the target audience including WHO, UNESCO, UNICEF and the UK and Scottish Governments. Findings from the review will be disseminated via an article in the Conversation, seminar and conference presentations, and podcasts posted on the project website and disseminated by social media.

## Discussion

Our review will be the first critical realist (or realist) review of the literature on mindfulness interventions in schools. In the review, we aim to identify the generative mechanisms and social structures that explain how and why school-based mindfulness interventions promote the mental wellbeing of CA. This will enable policymakers and school leaders to understand under what circumstances school-based mindfulness interventions promote CA mental wellbeing and for which CA they work.

The main challenges are likely to be that: most studies will have been carried out in high-income countries, so we will not be able to identify critical context elements and relevant mechanisms that are unique to low-and-middle-income countries; there will be little information in the documents on the context and; the documents may not include details of the theoretical reasoning underpinning their intervention to enable us to develop middle-range theories.

## Authors Contributions

PA, GN and LD contributed to the NIHR grant application. PA led on the design of the research and the writing of the protocol and produced the first draft. GN, IS and LD revised drafts and agreed the final text of this paper.

## Data Availability

No datasets were generated or analysed during the current study. All relevant data from this study will be made available upon study completion.

