## Additional Material 1 for "A protocol for a systematic critical realist synthesis of school mindfulness interventions designed to promote pupils’ mental wellbeing"

**PRISMA-P 2015 Checklist**

| **Section and topic** | **Item No** | **Checklist item** | **Page** |
| --- | --- | --- | --- |
| **ADMINISTRATIVE INFORMATION** | | | |
| Title: |  |  |  |
| Identification | 1a | The report is identified as a protocol of a critical realist synthesis review | 2 |
| Update | 1b | The protocol is not an update of a previous systematic review | n/a |
| Registration | 2 | The review is registered with PROSPERO | 3 |
| Authors: |  |  |  |
| Contact | 3a | The name, institutional affiliation, and e-mail address of all protocol authors are provided with the physical mailing address of the corresponding author | 1 |
| Contributions | 3b | The contributions of protocol authors are described | 24 |
| Amendments | 4 | The protocol is not an amendment of a previously completed or published protocol | n/a |
| Support: |  |  |  |
| Sources | 5a | Sources of financial support for the review have been identified | Financial disclosure |
| Sponsor | 5b | The review funder and sponsor have been provided | Financial disclosure |
| Role of sponsor or funder | 5c | The role of external support in developing the protocol has been provided | n/a |
| Rationale | 6 | The rationale for the review in the context of what is already known is described | 3 - 6 |
| Objectives | 7 | An explicit statement of the question(s) the review will address with reference to participants, interventions, comparators, and outcomes (PICO) is provided | 6 - 7 |
| Eligibility criteria | 8 | The study characteristics to be used as criteria for eligibility for the review are described in accordance with realist review publication standards | 14 |
| Information sources | 9 | All intended information sources are described | 12 - 14 |
| Search strategy | 10 | The search strategy is described | 12 - 14 |
| Study records: |  |  |  |
| Data management | 11a | The tools that will be used to manage records and data throughout the review are described | 14 |
| Selection process | 11b | The study selection process is described, including the involvement of a second reviewer to corroborate the data extracted and theory generation | 15 - 16 |
| Data collection process | 11c | The planned method of extracting data from reports is provided | 15 -16 |
| Data items | 12 | The data items relate to the context, agency, intervention, and mechanisms configuration, which has been described in detail | 16-18 |
| Outcomes and prioritisation | 13 | The outcomes for which data will be sought relate to SBMIs as described within the protocol. There is no prioritisation | 16 - 18 |
| Risk of bias in individual studies | 14 | n/a as critical realist reviews consider the relevance and rigour of included studies, this process is described | n/a |
| Data synthesis | 15a | The criteria under which study data will be synthesised are described | 18 |
|  | 15b | n/a data will not be extracted for quantitative synthesis | n/a |
|  | 15c | n/a data will not be extracted for additional analyses (such as sensitivity or subgroup analyses, meta-regression) | n/a |
|  | 15d | n/a data will not be extracted for quantitative synthesis; therefore, alternative synthesis methods are not appropriate | n/a |
| Meta-bias(es) | 16 | The realist review is not considering any planned assessment of meta-bias(es) (such as publication bias across studies, selective reporting within studies) | n/a |
| Confidence in cumulative evidence | 17 | The strength of the body of evidence will be determined using a series of judgements from the review authors in alignment with previous critical realist reviews and realist review publication standards | 7 |

Source: Moher D et al.: Preferred reporting items for systematic review and meta-analysis protocols (PRISMA-P) 2015 statement. Systematic Reviews 2015 4:1
